# Semaglutide Slows Epigenetic Aging in People with HIV-associated lipohypertrophy: Evidence from a Randomized Controlled Trial

**DOI:** 10.1101/2025.07.09.25331038

**Authors:** Michael J. Corley, Varun Dwaraka, Alina PS Pang, Danielle Labbato, Ryan Smith, Allison Ross Eckard, Grace A. McComsey

## Abstract

Semaglutide is a once-weekly GLP-1 receptor agonist that has been proposed as a gerotherapeutic, yet no data exist on its effects on epigenetic aging. We therefore conducted a post-hoc epigenetic analysis of a 32-week, double-blind, placebo-controlled phase 2b trial in adults with HIV-associated lipohypertrophy (semaglutide n = 45; placebo n = 39). Paired peripheral-blood methylomes were profiled to evaluate semaglutide’s impact across multiple generations of DNA-methylation clocks. After adjustment for sex, BMI, hsCRP, and sCD163, semaglutide significantly decreased epigenetic aging: PCGrimAge (-3.1 years, P = 0.007), GrimAge V1 (-1.4 years, P = 0.02), GrimAge V2 (-2.3 years, P = 0.009), PhenoAge (-4.9 years, P = 0.004), and DunedinPACE (-0.09 units, ≈9 % slower pace, P = 0.01). Semaglutide also lowered the multi-omic OMICmAge clock (-2.2 years, P = 0.009) and the transposable element-focused RetroAge clock (-2.2 years, P = 0.030). Eleven organ-system clocks showed concordant decreased with semaglutide, most prominently inflammation, brain and heart, whereas an Intrinsic Capacity epigenetic clock was unchanged (P = 0.31). These findings provide, to our knowledge, the first clinical-trial evidence that semaglutide modulates validated epigenetic biomarkers of aging, justifying further evaluation of GLP-1 receptor agonists for health-span extension.

## INTRODUCTION

Aging is the primary driver of chronic diseases, multimorbidity, and mortality worldwide, positioning interventions targeting biological aging as transformative therapeutic strategies with potential to substantially improve human healthspan(López-Otín et al. 2023; Kennedy et al. 2014). Within the emerging geroscience paradigm(Kennedy et al. 2014), pharmacologic agents originally developed for metabolic indications, such as glucagon-like peptide-1 (GLP-1) receptor agonists(Wilding et al. 2021; Davies et al. 2021; Marso et al. 2016; Maretty et al. 2025), have garnered attention due to their potential dual roles in metabolic regulation and aging biology. Semaglutide, a once-weekly glucagon-like peptide-1 (GLP-1) receptor agonist, has emerged as a transformative therapeutic for its marked weight reduction and cardiometabolic benefits, including reductions in visceral adipose tissue(Wilding et al. 2021; Davies et al. 2021; Marso et al. 2016). Given that obesity and adiposity embed an obesogenic epigenetic memory and are linked to accelerated epigenetic aging(de Toro-Martín et al. 2019; Lundgren et al. 2022; Hinte et al. 2024), there is growing interest in whether semaglutide’s metabolic effects might slow or reverse aspects of biological aging. Despite extensive evidence supporting semaglutide’s pleotropic benefits, randomized clinical trial data evaluating its effects on aging biomarkers remain absent.

People with HIV (PWH) represent a unique population exhibiting accelerated biological aging, characterized by premature onset of age-related conditions, persistent low-grade inflammation, and metabolic dysfunction, even when HIV replication is effectively suppressed by antiretroviral therapy (ART)(Deeks 2011; Breen et al. 2022; Pathai et al. 2014). A common metabolic complication in this population is HIV-associated lipohypertrophy, defined by excessive accumulation of visceral and ectopic adipose tissue, which further exacerbates aging processes(Koethe et al. 2020). Within the geroscience framework(Montano et al. 2022), the accelerated-aging phenotype in PWH provides an ideal clinical model to evaluate candidate geroprotective therapies, with findings potentially relevant to the general aging population. This setting may allow earlier insights into treatment effects, particularly via DNA-methylation based epigenetic clocks and other emerging aging biomarkers, which are recently being considered as outcome measures in double-blind, randomized geroscience trials(Kroemer et al. 2025) and global competitions testing gerotherapeutics(Justice 2024).

In a completed phase 2b, randomised, double-blind, placebo-controlled trial, we tested whether once-weekly semaglutide can slow epigenetic aging in people with HIV-associated lipohypertrophy, a population marked by visceral adiposity and accelerated epigenetic age. Using paired peripheral-blood methylomes collected at baseline and 32 weeks, we conducted a post-hoc analysis spanning 17 first-, second-, and third-generation DNA-methylation clocks. Here we test whether once-weekly semaglutide slows DNA-methylation aging in this high-risk population. To our knowledge, this provides the first randomized clinical evidence that a licensed GLP-1 receptor agonist modulates epigenetic biomarkers of aging, positioning semaglutide as a candidate gerotherapeutic and laying the groundwork for prospectively powered, mechanism-focused trials aimed at extending healthspan in populations vulnerable to accelerated aging.

## RESULTS

### Posthoc Epigenetic Age Analysis of a Randomized Trial of Semaglutide

To determine whether semaglutide treatment could impact biological aging, we conducted a post hoc epigenetic analysis of participants enrolled in a previously reported 32-week, randomized, double-blind, placebo-controlled phase 2b clinical trial evaluating semaglutide in people with HIV (PWH) and lipohypertrophy (Eckard et al. 2024)(**Figure 1**). The trial aimed to evaluate the effects of the GLP-1 receptor agonist semaglutide on adipose tissue quantity and distribution in individuals with HIV-associated lipohypertrophy over a 32-week period. Eligible participants included adults aged 18 years or older with documented HIV-1 infection, stable ART for at least 12 weeks, and controlled HIV-1 RNA levels (<400 copies per mL) for six months prior to screening. Additional inclusion criteria were a BMI of 25 kg/m² or greater and the presence of lipohypertrophy without type 1 or type 2 diabetes or cardiovascular disease. Participants were randomly assigned 1:1 to receive either once-weekly subcutaneous semaglutide (8-week dose titration followed by 24 weeks at 1.0 mg) or matching placebo. All weekly injections were provided in the clinic by a certified nurse. Randomization was performed using an online software program with block sizes of six, and treatment assignments were masked to participants, investigators, and research personnel. Of 154 individuals assessed for eligibility, 108 were randomized (54 in each group). Eight participants (15%) from each group withdrew prematurely. The study’s primary outcomes included changes in adipose tissue quantity across body compartments; abdominal visceral and subcutaneous adipose tissue were measured by L4-L5 non contrast abdominal CT scan imaging, while total body fat, trunk and peripheral fat were measured by whole body DEXA scan. Secondary outcomes included metabolic measures (glucose metabolism, insulin resistance, lipid profiles), anthropometric changes (weight, BMI, waist-to-hip ratio), inflammation and immune activation markers, and safety. The trial is described in more detail at ClinicalTrials.gov (NCT04019197) and in Methods. Results for the primary outcomes were published(Eckard et al. 2024). Peripheral blood mononuclear cells (PBMCs) were collected at baseline and 32 weeks follow-up and biobanked.

**Figure 1:**
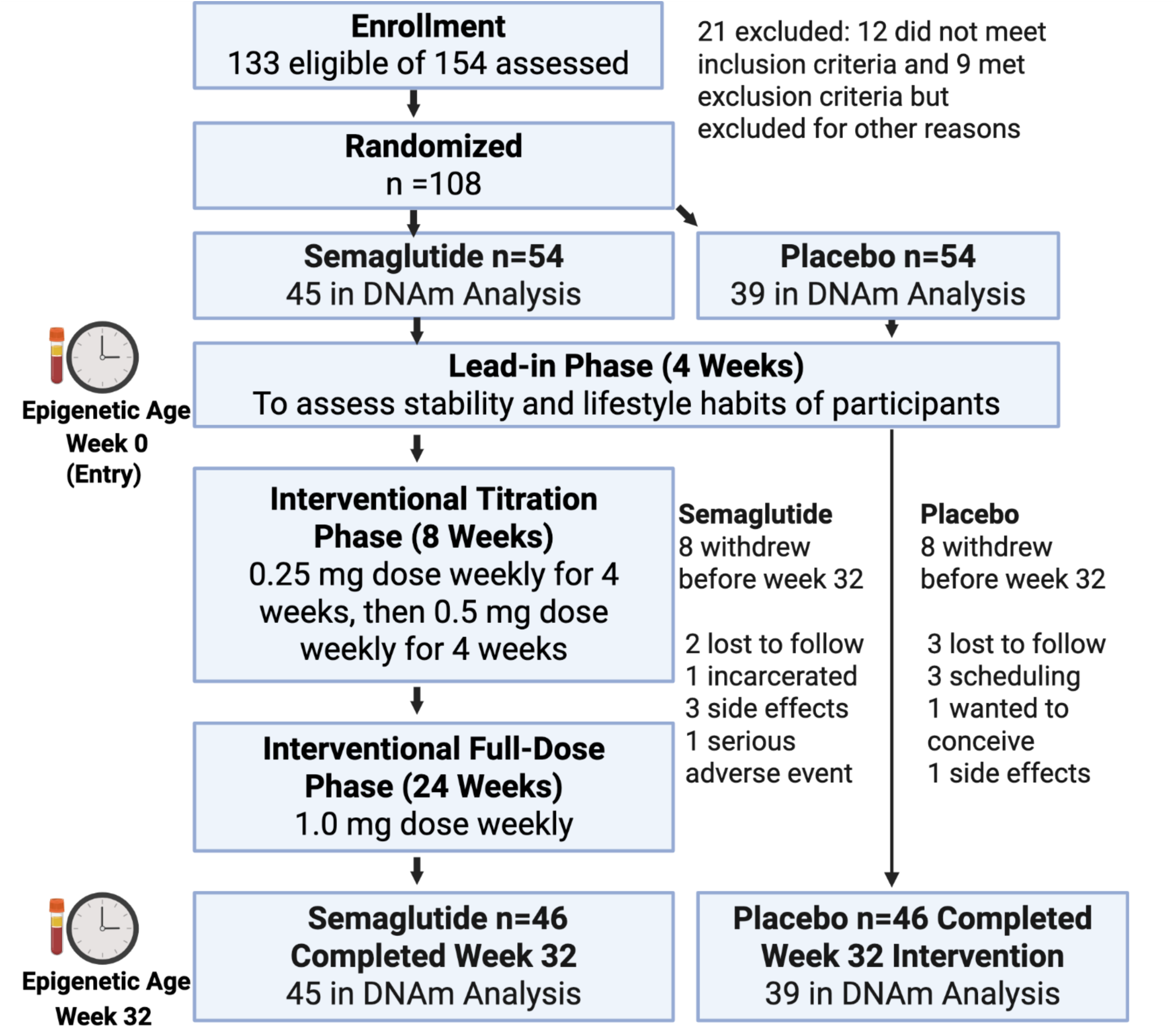
Consort Diagram for the semaglutide trial in PWH. Epigenetic age was assayed from peripheral blood mononuclear cell samples collected at baseline (Week 0) and the post interventional phase follow up time point (Week 32). Of the total 92 participants that completed longitudinal Week 0 and week 32 assessments, epigenetic age was assayed longitudinally for n = 45 semaglutide and n = 39 placebo participants.

We first quantified biological age using established first-(Horvath 2013; Hannum et al. 2013), second-(Levine et al. 2018; Lu et al. 2019), and third-generation epigenetic clocks(Belsky et al. 2022; Ying et al. 2024) in biobanked longitudinal peripheral-blood mononuclear cells from 84 total participants (45 semaglutide, 39 placebo) using epigenetic DNA-methylation data obtained at baseline and week 32. We also utilized versions of 1^st^ and 2^nd^ generation clocks built from DNAm principal components (PCs) (termed ‘PC clocks’) that show enhanced technical reliability and utility in longitudinal study designs such as randomized clinical trials(Higgins-Chen et al. 2022).

For the epigenetic analysis, at baseline the 84 participants (45 semaglutide, 39 placebo) were middle-aged, with a mean ± SD age of 49 ± 12 years and well balanced between treatment arms (48 ± 13 vs. 50 ± 12 years). 42% were women overall, but men were slightly over-represented in the semaglutide group (67 % vs. 49 %). The cohort included 58 % Black, 38 % White and 11 % Hispanic participants with nearly identical distributions across groups. Immunologically, CD4 counts were high (median 762 cells µL⁻¹) and CD4/CD8 ratios near 1.0, reflecting immune reconstitution; nadir CD4 counts were lower, as expected, but similar between arms. Viral suppression was durable: only 9 % had HIV-1 RNA above the lower limit of quantification, and ART duration averaged ∼14 years. Participants were obese (median BMI 32.9 kg m⁻²) with comparable anthropometry in each group. One-third were current smokers, another quarter former smokers. Glycemic control was normal (median HbA1c 5.5 %), although insulin resistance was evident (median fasting HOMA-IR 2.9), again without meaningful group differences. Estimated 10-year ASCVD risk was moderate at 4.7 % (IQR 2.2–8.0). Inflammatory biomarkers showed low-grade activation: median high-sensitivity CRP 4.1 µg/mL and sCD163 605 pg mL⁻¹, with slightly higher values in placebo. Overall, baseline characteristics of participants assayed in the epigenetic analysis were well matched. (**Table 1**).

**Table 1:** Baseline characteristics of the clinical trial population overall and by treatment group.

| Baseline Characteristic |  | All participants<br>(N = 84) | Semaglutide (n=45) | Placebo (n=39) |
| --- | --- | --- | --- | --- |
| Age (years) | Mean (SD) | 49 (12.5) | 48 (13.1) | 50 (11.8) |
| Sex | Male | 49 (58%) | 30 (67%) | 19 (49%) |
|  | Female | 35 (42%) | 15 (33%) | 20 (51%) |
| Race or ethnicity | Black | 49 (58%) | 26 (58%) | 23 (59%) |
|  | White | 32 (38%) | 18 (40%) | 14 (36%) |
|  | Biracial | 2 (2%) | 0 | 2 (5%) |
|  | Native American | 1 (1%) | 1 (2%) | 0 |
|  | Hispanic | 9 (11%) | 4 (9%) | 5 (13%) |
| HIV Variables | CD4 T-cell count, cells per ul | 762 (525, 1058) | 830 (389.3, 1081) | 728 (547, 923) |
|  | CD8 T-cell count, cells per ul | 806 (571, 1054) | 762.5 (572, 1074) | 847 (571, 1023) |
|  | CD4/CD8 Ratio | 0.98 (0.61, 1.43) | 1.04 (0.51, 1.31) | 0.97 (0.61, 1.43) |
|  | Nadir CD4, cells per ul | 211 (97, 377) | 266 (105.5, 509) | 163 (62.25, 313) |
|  | HIV-1 RNA (copies/mL) >LLQ | 8 (9%) | 3 (7%) | 5 (13%) |
|  | HIV duration, months | 228.8 (139.7, 312.6) | 200 (113.4, 491.2) | 235.8 (158.4, 301.7) |
|  | Antiretroviral therapy duration, months | 175.1 (107.5, 223.8) | 184.6 (97.49, 223.2) | 160.5 (109.9, 225.6) |
| Anthropometric Measurements | BMI, kg/m<sup>2</sup> | 32.86 (28.82, 38.97) | 32.80 (28.61, 35.93) | 33.09 (28.99, 39.73) |
| Lifestyle variables | Current Smoker | 29 (35%) | 13 (29%) | 16 (41%) |
|  | Past Smoker | 21 (25%) | 12 (27%) | 9 (23%) |
|  | Never Smoked | 34 (40%) | 20 (44%) | 14 (36%) |
| Glucose Metabolism and Insulin Resistance | HbA1c % | 5.5 (5.2, 5.8) | 5.4 (5.1, 5.8) | 5.6 (5.4, 5.9) |
|  | Fasting HOMA-IR | 2.9 (2.0, 5.1) | 2.6 (1.4, 4.6) | 3.7 (2.2, 7.5) |
|  | 2-h OGTT HOMA-IR | 10.3 (5.1, 26.8) | 9.9 (4.9, 19.1) | 10.4 (5.1, 42.0) |
| Cardiovascular Disease Risk | 10-year atherosclerotic cardiovascular disease risk estimate, % | 4.7 (2.2, 8.0) | 4.5 (2.4, 7.7) | 4.8 (1.9, 8.8) |
| Plasma Biomarkers | Hs-CRP (µg/mL) | 4.142 (2.021, 7.976) | 3.785 (1.743, 6829) | 5.713 (2.663, 9.668) |
|  | ss (pg/L) | 604.7 (456.4, 849.1) | 591.9 (458.1, 752.8) | 616.9 (446.9, 993.7) |
|  | sCD14 (pg/L) | 1785 (1505, 2122) | 1683 (1398, 2085) | 1927 (1650, 2130) |

### Semaglutide Slows Epigenetic Aging Across Multiple DNA Methylation Clocks

We evaluated the impact of semaglutide treatment on biological aging using 17 DNA methylation (DNAm)-based epigenetic clocks spanning first-, second-, and third-generation models. In ANCOVA models adjusting for baseline covariates (sex, BMI, hsCRP, and sCD163), semaglutide was associated with significantly reduced epigenetic aging relative to placebo across multiple clocks (**Figure 2A**). The most pronounced effects were observed for second-, and third-generation epigenetic clocks. Specifically, for PCGrimAge (a mortality-risk DNAm age estimate), semaglutide was associated with a 3.08-year lower annual epigenetic age increase compared to placebo (ANCOVA adjusted difference = –3.08 years/year, 95% CI: –5.29 to –0.86, p = 0.007). For DunedinPACE, semaglutide was associated with a 0.09 lower pace-of-aging (units per year) relative to placebo (95% CI: –0.17 to –0.02, p = 0.01). These estimates translate to roughly a 9% slower pace of aging and a 3-year reduction in annual biological age increase in the semaglutide group compared to placebo. Semaglutide also significantly reduced biological age acceleration in SystemsAge (–4.17 years/year, *p* = 0.009), PhenoAge (–4.90 years/year, *p* = 0.004), PCPhenoAge (–3.68 years/year, *p* = 0.02), and OMICmAge (–2.20 years/year, *p* = 0.009) (Chen et al. 2023) a next-generation DNA-methylation clock derived by modelling an electronic-medical-record–based aging phenotype with epigenetic, proteomic, metabolomic and clinical biomarker proxies. RetroClock, an epigenetic clock designed to capture retrotransposon-associated aging(Ndhlovu et al. 2024), also showed a significant response to semaglutide treatment. In adjusted models, semaglutide was associated with a –2.18 year/year reduction in RetroClock age compared to placebo (95% CI: –4.14 to –0.21; *p* = 0.030). In contrast to the more robust effects observed across mortality- and systems-level clocks, AdaptAge, CausAge, and DamAge (Ying et al. 2024) exhibited modest, directionally heterogeneous, and statistically non-significant changes with semaglutide treatment. AdaptAge increased by 3.49 years (95% CI: –2.20 to 9.18; *p* = 0.23), CausAge by 0.46 years (95% CI: –2.40 to 3.32; *p* = 0.75), and DamAge decreased by –2.22 years (95% CI: –7.09 to 2.65; *p* = 0.37) compared to placebo, highlighting the differential responsiveness of epigenetic aging clocks.

**Figure 2.**
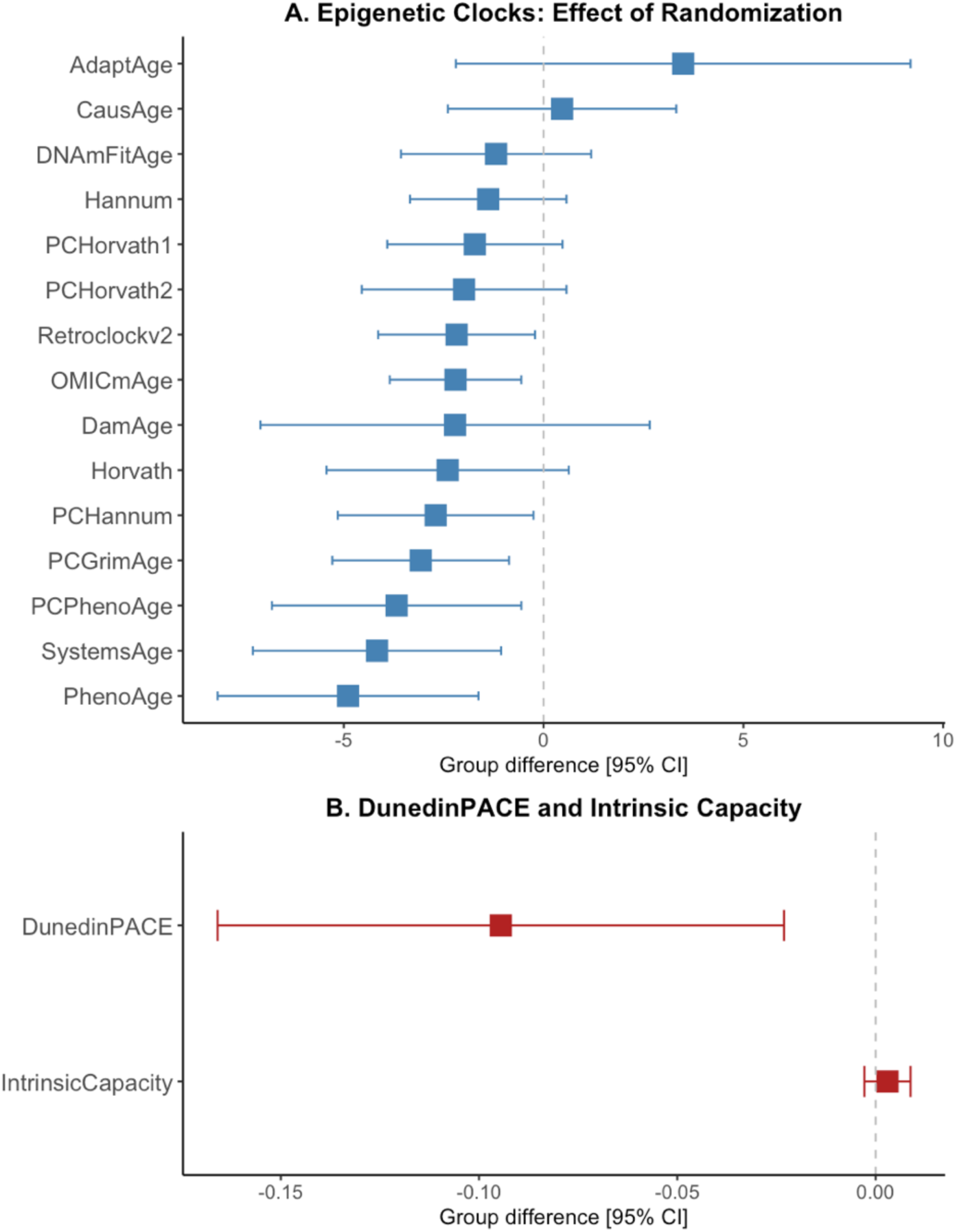
Semaglutide broadly decelerates epigenetic aging across multiple methylation clocks. Forest plot displaying the estimated difference (semaglutide − placebo) in annualized epigenetic age change for DNA methylation–based biomarkers of aging. Estimates are derived from ANCOVA models adjusting for baseline age, sex, BMI, and inflammation (HsCRP and sCD163), with bars indicating 95% confidence intervals. **(A)** Effects on 15 clocks spanning classical (Horvath, Hannum), principal component–based (PCHorvath1/2, PCHannum, PCPhenoAge), and newer modeling frameworks (PhenoAge, PCGrimAge, AdaptAge, CausAge, DamAge, DNAmFitAge, OMICmAge, Retroclockv2, SystemsAge). **(B)** DunedinPACE, a continuous pace-of-aging measure, and Intrinsic Capacity epigenetic clock. Negative estimates indicate slower biological aging in the semaglutide group. Semaglutide treatment was associated with a consistent deceleration across nearly all measures, highlighting its geroprotective potential.

### Intrinsic Capacity Clock Shows No Significant Change with Semaglutide Treatment

A DNA methylation-based epigenetic clock of Intrinsic Capcity (IC) was recently developed using elastic net regression in the INSPIRE-T cohort, identifying 91 CpG sites predictive of IC scores derived from clinical assessments(Fuentealba et al. 2025). This IC epigenetic clock showed strong age-related decline and only modest overlap with traditional epigenetic clocks, suggesting it captures a distinct axis of biological aging linked to physical and cognitive resilience. Hence, we evaluated the responsiveness of this first-generation Intrinsic Capacity (IC) clock to semaglutide in our study. In ANCOVA models adjusting for baseline age, sex, BMI, high-sensitivity CRP, and sCD163, semaglutide treatment did not significantly alter IC clock estimates over the 32-week period (estimated group difference: 0.003 IC score; *p* = 0.31) (**Figure 2B**). Caution is warranted in responsiveness of first-generation IC epigenetic clock changes to semaglutide and warrants further study.

### Semaglutide reduction in epigenetic age as measured by GrimAge and GrimAgeV2 using Biolearn

As an orthogonal computational approach to assess biological aging with second-generation mortality-based epigenetic clocks, we used Biolearn, an open-source library for biomarkers of aging, to examine whether semaglutide treatment significantly impacted DNA methylation-based age estimates for GrimAge V1(Lu et al. 2019) and the updated GrimAge V2(Ying et al. 2023; Lu et al. 2022). Using adjusted ANCOVA models controlling for BMI, high-sensitivity C-reactive protein (HsCRP), soluble CD163, and sex, we found that randomization to semaglutide was significantly associated with reductions in both GrimAge clocks. Participants randomized to semaglutide exhibited a 1.39-year lower GrimAgeV1 estimate compared to placebo (95% CI: –2.72 to –0.05; *p* = 0.042) over the 32-week trial. The effect was more pronounced in GrimAgeV2, with a 2.26-year reduction relative to placebo (95% CI: –3.94 to – 0.59; *p* = 0.008). These findings, derived using the Biolearn modeling framework, provide orthogonal confirmation that semaglutide may decelerate biological aging as captured by mortality risk–associated epigenetic signatures.

### Semaglutide Broadly Decelerates Multi-system Epigenetic Aging Across 11 System Clocks

Because GLP-1 receptor agonists like semaglutide have demonstrated pleiotropic benefits that span cardiometabolic, renal, hepatic and neuroprotective domains(Zheng et al. 2024), we next evaluated whether these effects extended to organ-specific biological aging. We applied a panel of 11 DNA methylation–based “system clocks” derived from a single blood methylation assay that deconvolves biological aging across individual organ systems, including blood, brain, inflammation, heart, hormone, immune, kidney, liver, metabolic, lung, and musculoskeletal domains(Sehgal et al. 2023). Each system clock captures both all-cause mortality risk and organ-specific decline; for example, the Heart clock predicts cardiovascular events, while the Brain clock tracks cognitive function and neuroimaging correlates. In adjusted ANCOVA models controlling for age, sex, BMI, hsCRP, and sCD163, semaglutide treatment was associated with consistent reductions in epigenetic age across all 11 systems (**Figure 4**). The largest effects were observed in the Blood (–4.37 years, *p* = 0.011), Brain (–4.99 years, *p* = 0.0049), and Inflammation (–5.01 years, *p* = 0.0056) clocks. Substantial deceleration was also observed in the Heart (–4.34 years, *p* = 0.0088), Kidney (–4.20 years, *p* = 0.014), Liver (–4.19 years, *p* = 0.042), and Metabolic (–4.72 years, *p* = 0.0090) clocks. While trends were directionally favorable, semaglutide-related reductions in epigenetic age did not reach statistical significance for the Lung (–2.21 years; 95% CI: –5.19 to 0.76; *p* = 0.14), Hormone (–1.33 years; 95% CI: – 4.01 to 1.35; *p* = 0.33), Immune (–1.60 years; 95% CI: –4.83 to 1.64; *p* = 0.33), and Musculoskeletal (–2.32 years; 95% CI: –5.45 to 0.81; *p* = 0.15) system clocksThese findings highlight the potential of semaglutide to exert geroprotective effects that extend across multiple physiological systems.

**Figure 3.**
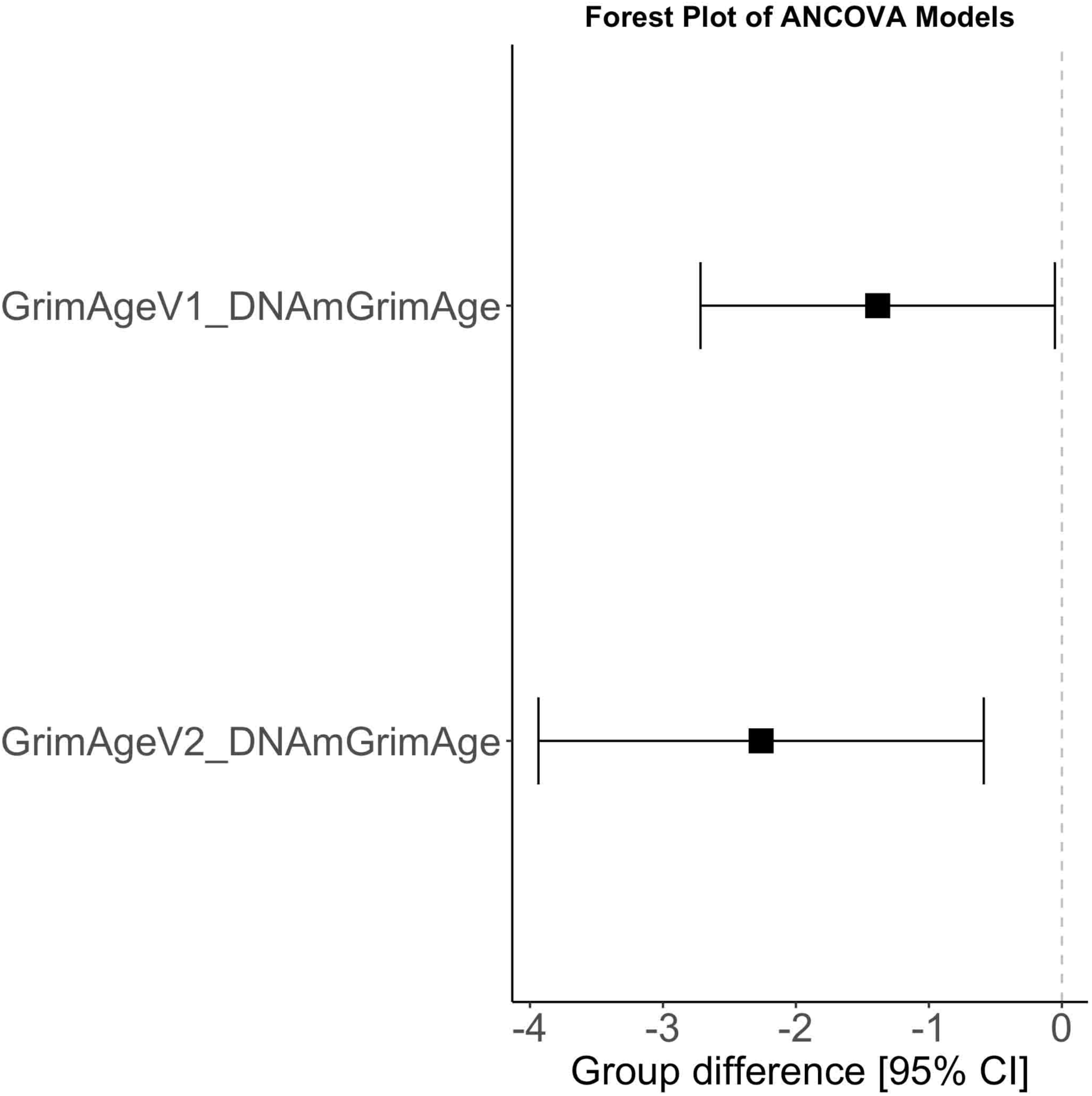
Forest plot showing the effect of semaglutide on GrimAge V1 and V2 epigenetic aging measures calculated from Biolearn. Adjusted ANCOVA models were used to assess the association between semaglutide treatment (Randomization) and DNA methylation-based aging estimates, GrimAgeV1_DNAmGrimAge and GrimAgeV2_DNAmGrimAge, while controlling for BMI, high-sensitivity C-reactive protein (HsCRP), soluble CD163, and sex. Estimates represent the mean difference in years between treatment and control groups, with 95% confidence intervals. Both GrimAgeV1 and GrimAgeV2 showed significant reductions in epigenetic age in the semaglutide group, with a stronger effect observed for GrimAgeV2 (*p* = 0.0088).

**Figure 4.**
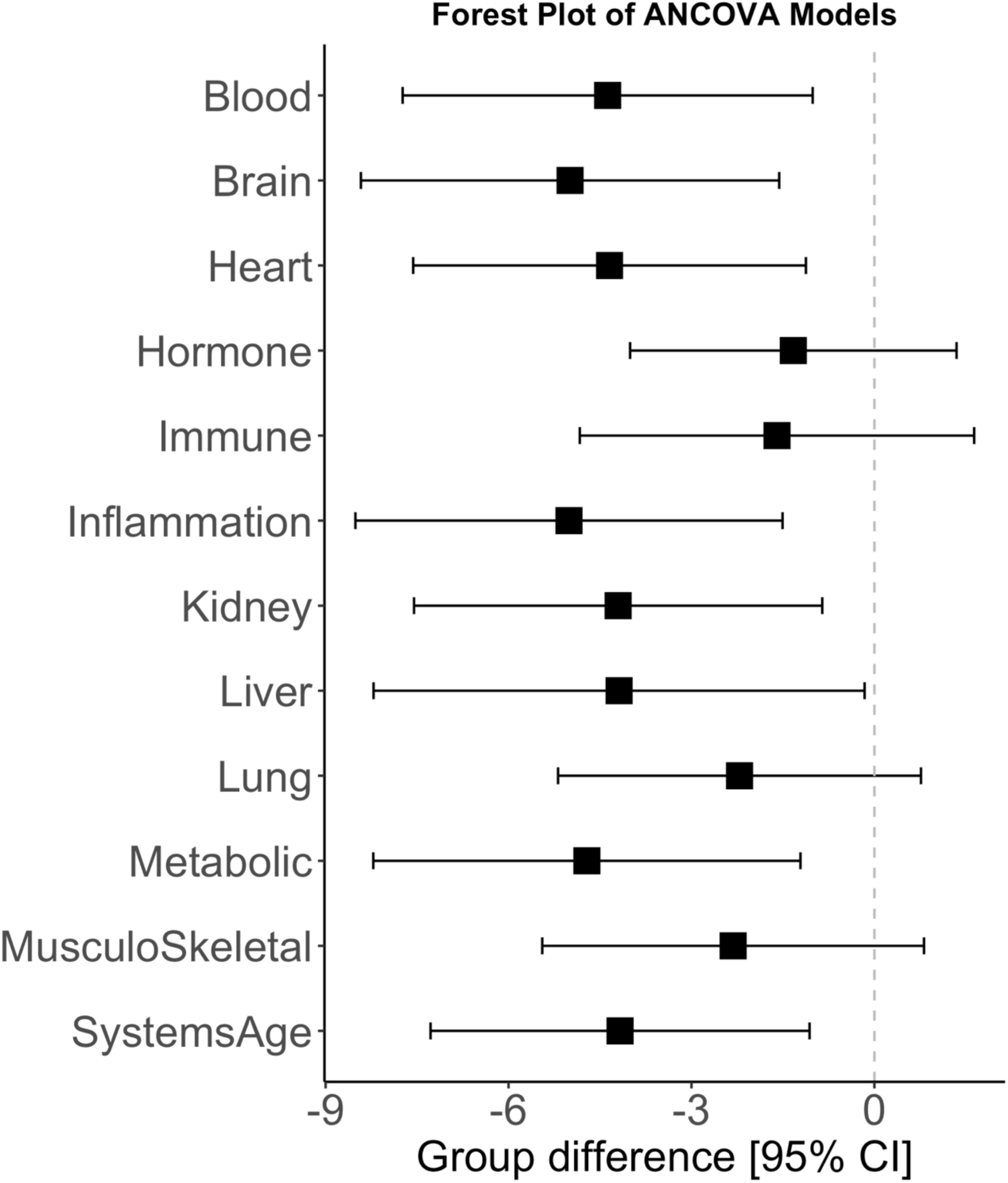
Semaglutide confers a multi-organ deceleration of system-level epigenetic aging. Forest plot of the between-group difference (semaglutide − placebo) in annualized epigenetic age (years yr⁻¹) across 11 organ-system clocks (Blood, Brain, Heart, Hormone, Immune, Inflammation, Kidney, Liver, Lung, Metabolic, Musculoskeletal) plus a composite SystemsAge index. Data points represent adjusted mean differences (± 95% confidence intervals) estimated by ANCOVA, controlling for baseline age, sex, body-mass index, and inflammation. Negative values to the left of the dashed line indicate slower system-specific aging in the semaglutide group, with the largest effects seen in Blood, Brain, and Inflammation (∼ –6 years). The SystemsAge aggregate measure was ∼ –4 years, underscoring a broad, multi-organ geroprotective signal.

## Discussion

This study provides, to our knowledge, the first randomized clinical trial evidence that once-weekly semaglutide can modulate biological aging, as measured by DNA methylation-based epigenetic clocks. Over the 32-week intervention period in individuals with HIV-associated lipohypertrophy, a population characterized by metabolic dysfunction and accelerated biological aging, semaglutide treatment led to robust attenuation and, in some cases, reversal of age-related DNA methylation signatures. Notably, the most pronounced effects were observed in second-generation clocks predictive of morbidity and mortality risk and in third-generation measures of biological aging rate DunedinPACE) suggesting that semaglutide may exert pleiotropic effects that extend beyond metabolic regulation to influence biological aging.

Our results align well with emerging geroscience paradigms, which propose that lifestyle and metabolic interventions capable of reducing adiposity, systemic inflammation, and insulin resistance could significantly modulate aging trajectories and improve healthspan(Kennedy et al. 2014). Notably, similar epigenetic clock signals have been observed with caloric restriction (CALERIE trial; (Waziry et al. 2023)) and multimodal lifestyle interventions (DO-HEALTH trial; (Bischoff-Ferrari et al. 2025)), supporting the broader concept that targeted metabolic modulation can impact aging pathways. Given the considerable burden of multimorbidity and mortality driven by age-related chronic conditions worldwide, identifying pharmacological agents like semaglutide capable of slowing biological aging represents a critical advancement for geroscience(Kroemer et al. 2025). Further longitudinal studies specifically examining the effects of GLP-1 receptor agonists on validated biomarkers of aging are warranted to clarify their role within the expanding geroscience therapeutic landscape.

Semaglutide’s multi-organ impact(Marso et al. 2016; Wilding et al. 2021; Badve et al. 2025; Davies et al. 2021), observed consistently across system-specific epigenetic clocks, further underscores its gerotherapeutic potential. Strong reductions in biological age within inflammation, brain, cardiovascular, hepatic, and renal systems suggest pleiotropic geroprotective mechanisms consistent with emerging literature indicating broad systemic benefits of GLP-1 receptor agonists. Indeed, we observed previously that semaglutide treatment markedly decreased inflammation-associated biomarkers (IL-6, sCD163) previously linked to morbidity and mortality in HIV (Funderburg et al. 2025). These findings align with emerging mechanistic studies in murine models showing that GLP-1R agonists can reverse obesogenic memory through anti-inflammatory and metabolic reprogramming of adipose tissue, targeting pathways such as CCL2/CCR2 that drive epigenetic and metabolic dysfunction(Léon et al. 2025). Recent proteomic studies have also revealed that semaglutide alters circulating proteins involved in lipid metabolism, inflammation, and cardiovascular risk pathways—independent of weight loss—suggesting broader reprogramming effects that could synergize with epigenetic aging deceleration(Maretty et al. 2025). These data suggest a plausible mechanistic link wherein semaglutide mitigates chronic immune activation and inflammatory signaling pathways central to aging acceleration, particularly relevant in populations experiencing chronic inflammation such as PWH.

Our results provide insights into potential biological mechanisms underlying semaglutide’s geroprotective effects. Given that epigenetic age acceleration correlated more closely with measures of central adiposity rather than systemic inflammation, semaglutide’s marked ability to reduce visceral adipose tissue may directly mitigate adipose-driven pro-aging signals such as dysregulated adipokine secretion and local inflammation. Recent studies have demonstrated that adipose tissue retains a persistent “obesogenic epigenetic memory,” characterized by stable transcriptional and chromatin accessibility changes even after significant weight loss, predisposing individuals to adverse metabolic responses upon weight regain(Hinte et al. 2024). Improved adipose tissue function and enhanced insulin sensitivity achieved through semaglutide treatment could thus disrupt or partially reverse these detrimental epigenetic signatures. Further mechanistic investigations into adipocyte-specific methylation patterns, chromatin remodeling, and transcriptional responses to semaglutide will be critical to confirm its role in counteracting adipose-derived epigenetic memory and reducing susceptibility to metabolic dysfunction and accelerated biological aging.

Our study also highlights important methodological insights regarding epigenetic clocks as outcome measures(Conole et al. 2025; Teschendorff & Horvath 2025; Perri et al. 2025). We observed heterogeneous responsiveness across different classes of epigenetic biomarkers. While mortality-linked clocks, pace of aging, and multi-system indices demonstrated robust treatment effects, clocks designed to capture resilience (AdaptAge, IC clock) or causally-driven aging (CausAge, DamAge) were less responsive. This finding emphasizes the need for careful biomarker selection in geroscience trials, acknowledging unique epigenetic clock-specific attributes. Moreover, despite these promising findings, several limitations of our findings should be considered. This study represents a post hoc analysis, limited by sample size and short follow-up duration. Longer-term trials with larger cohorts are necessary to validate durability, translate epigenetic changes into clinical outcomes, and assess generalizability beyond the unique HIV-associated lipohypertrophy population. Additionally, exploring alternative GLP-1 agonists, dosing or treatment regimens (e.g., microdosing) could enhance applicability to broader populations interested in preventive aging interventions.

To our knowledge, this study provides the first placebo-controlled randomized clinical trial evidence that a GLP-1 receptor agonist can modulate DNA methylation-based biomarkers of biological aging. These findings strongly support semaglutide’s potential as a gerotherapeutic capable of influencing fundamental aging processes. By demonstrating semaglutide’s robust impact across multiple validated epigenetic aging clocks, our results provide compelling rationale for systematically evaluating GLP-1 receptor agonists through FDA-approved drug repurposing frameworks, accelerating their clinical translation as geroscience-guided therapies to enhance healthspan and prevent chronic age-related diseases.

---

**Supplemental Figure 1:**
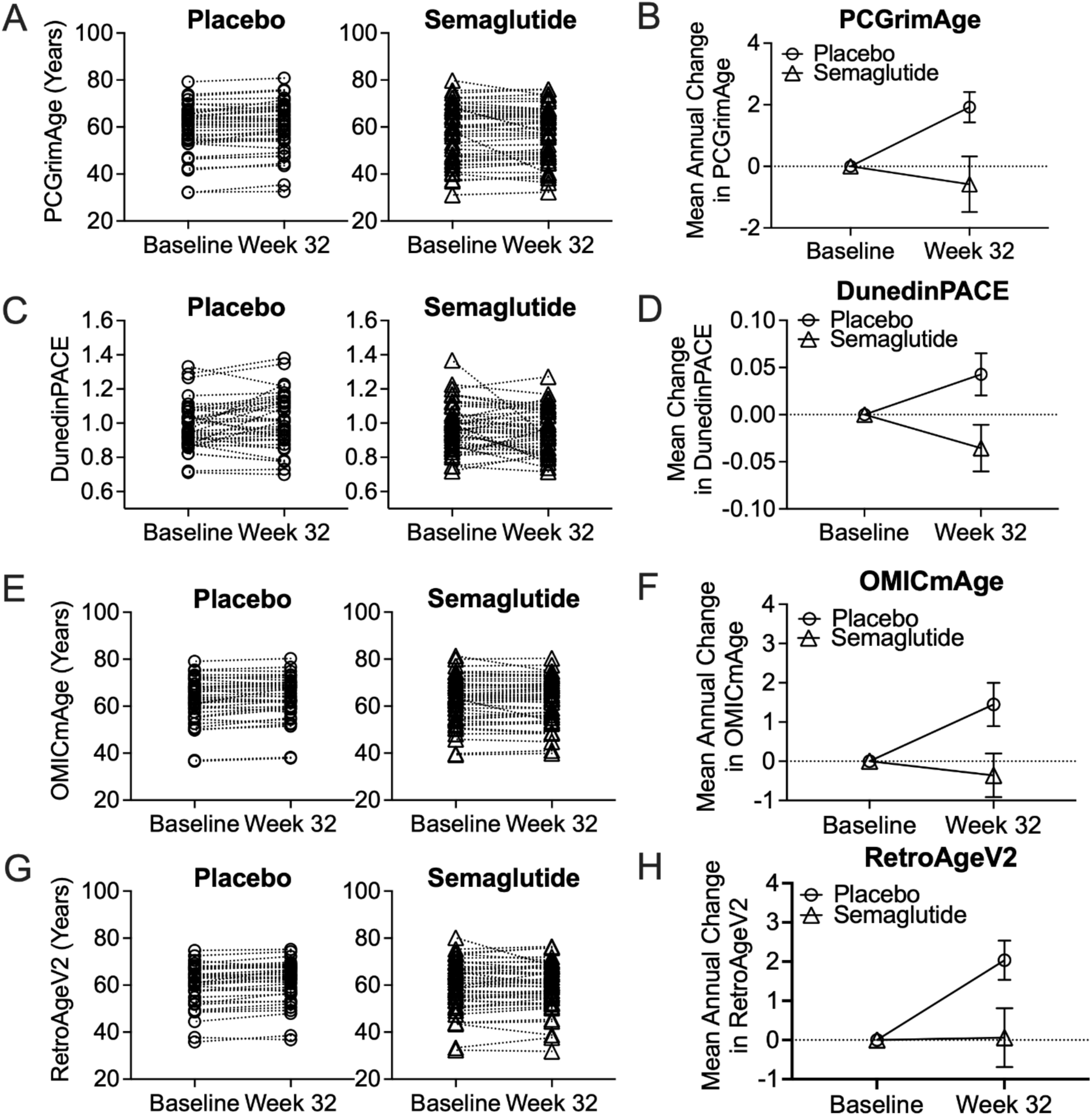
Change in epigenetic age from baseline to week 32 follow-up in PCGrimAge, DunedinPACE, OMICmAge, and RetroAgeV2 in the placebo and semaglutide groups. Figure shows semaglutide treatment effects on two well established epigenetic clock algorithms previously utilized to evaluate clinical trials effects on biological aging, PCGrimAge and DunedinPACE. The new epigenetic clocks OMICmAge and RetroAgeV2 are also presented. Calculated values are displayed in open circles for each participant in the placebo (n=39) and open triangles for semaglutide (n=45) group. Values for PCGrimAge, OMICmAge, and RetroAgeV2 are in years and DunedinPACE are in pace-of-aging units where the reference norm is 1.0. The right column shows the mean annual change from baseline.

**Supplemental Figure 2:**
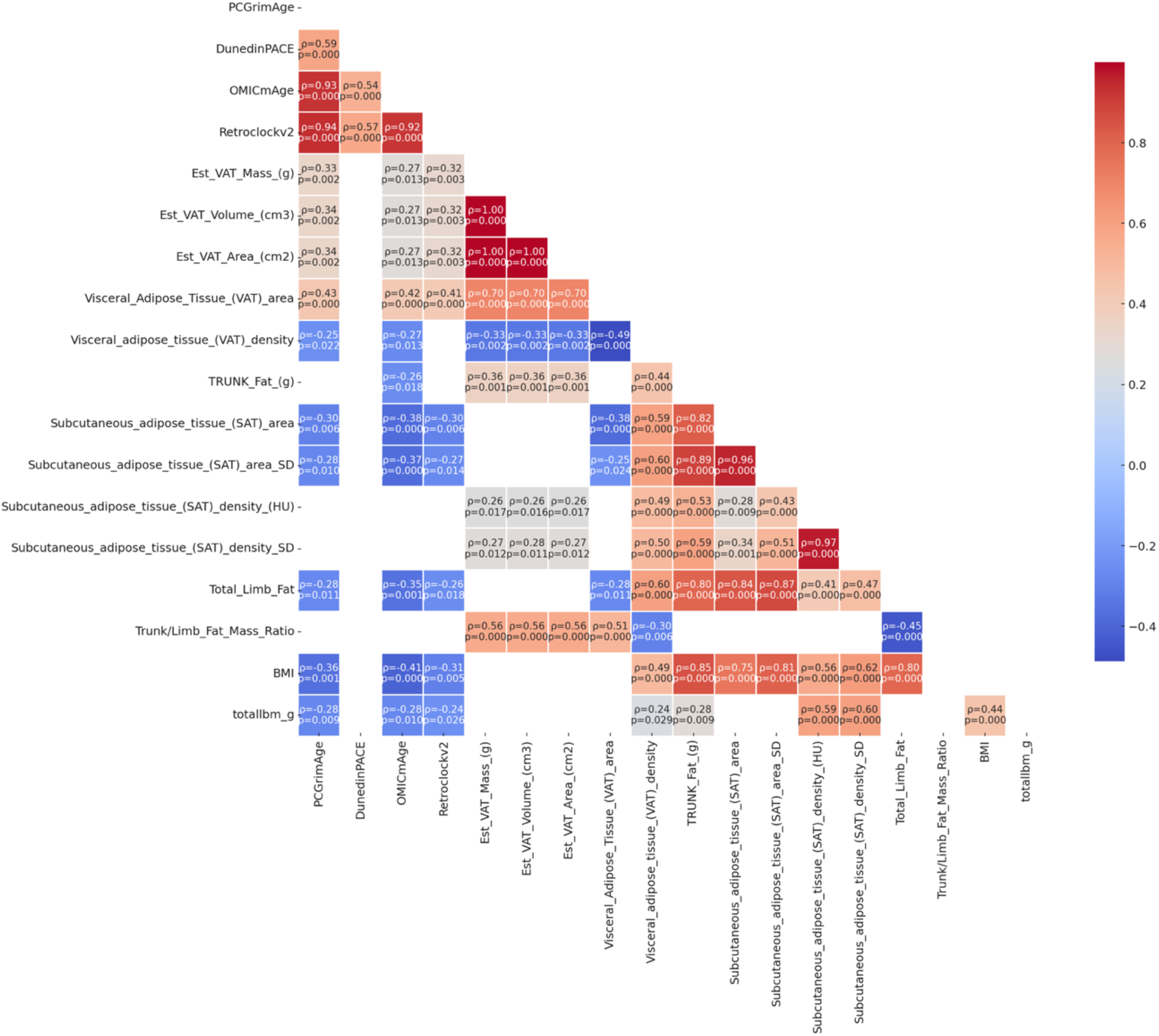
Epigenetic Age and Adiposity. Spearman correlation plot illustrating associations between epigenetic aging biomarkers and comprehensive adiposity phenotypes at baseline. Epigenetic aging measures include PCGrimAge, DunedinPACE, OMICmAge, and Retroclockv2. Adiposity was comprehensively assessed using visceral adipose tissue (VAT) measures (estimated mass, volume, area, and density), trunk fat, subcutaneous adipose tissue (SAT) characteristics (area, area standard deviation, density, density standard deviation), total limb fat, trunk-to-limb fat mass ratio, body mass index (BMI), and total body lean mass (totallbm_g). Spearman rho values are shown in colored boxes, with red indicating positive correlations and blue indicating negative correlations. Statistical significance (p-values) is indicated numerically within each cell, with significant correlations (p<0.05) highlighted by the intensity of color shading. Non-significant associations are shown white boxes.

**Supplemental Figure 3:**
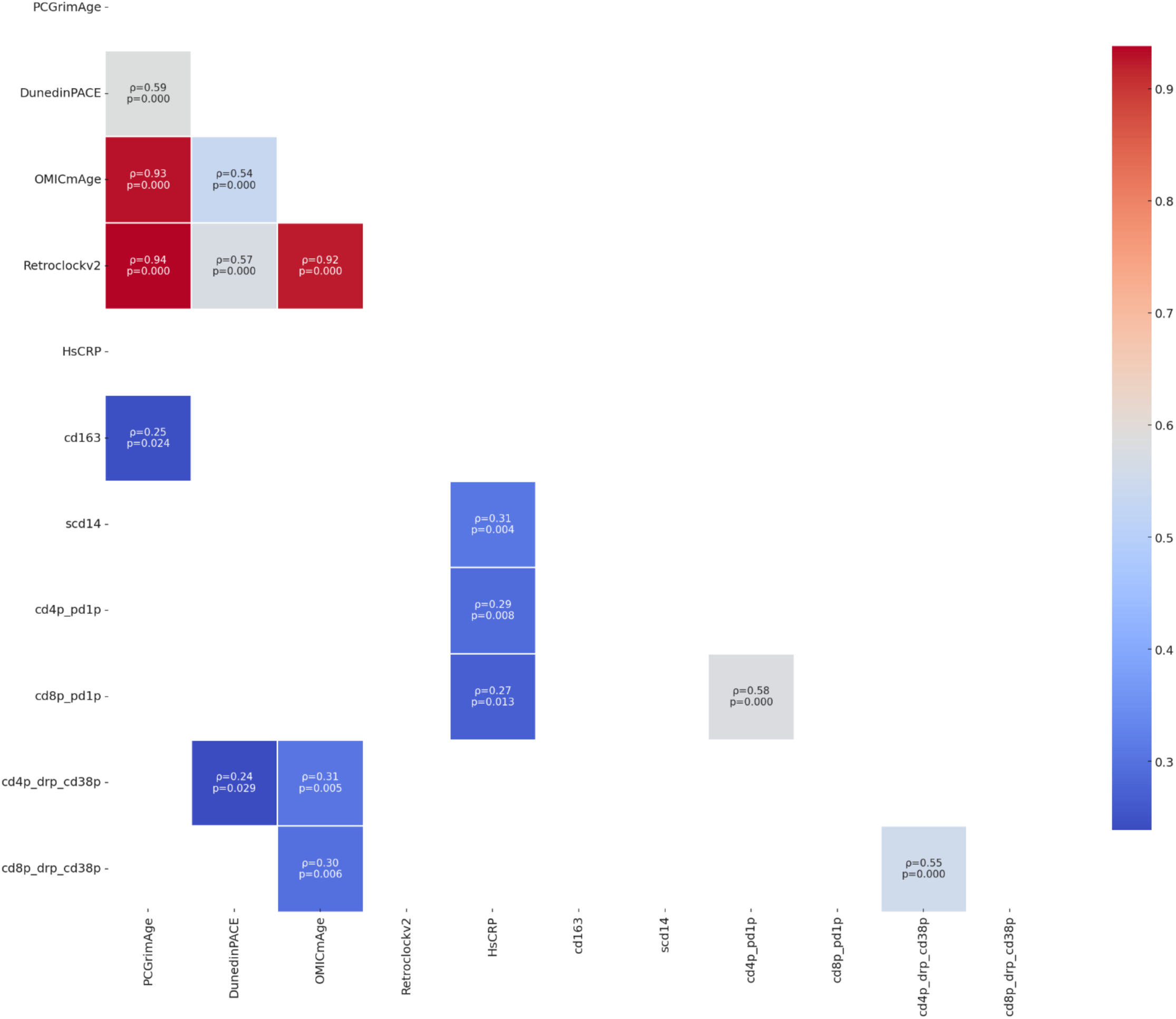
Epigenetic Age and Immune Biomarkers. The plot displays pairwise Spearman correlations (ρ) between epigenetic aging biomarkers (PCGrimAge, DunedinPACE, OMICmAge, Retroclockv2) and plasma inflammatory markers (HsCRP, CD163, sCD14) as well as flow cytometry–quantified T-cell subsets (PD-1⁺ CD4⁺ and CD8⁺ T cells, CD4⁺DR⁺CD38⁺, and CD8⁺DR⁺CD38⁺). Only statistically significant correlations (*p* < 0.05) are shown. Each annotated cell includes the correlation coefficient (ρ) and corresponding *p*-value. Strongest associations were observed between OMICmAge and both CD4⁺DR⁺CD38⁺ and CD8⁺DR⁺CD38⁺ T-cell subsets. Gray cells indicate non-significant correlations (*p* ≥ 0.05).

## Materials and Methods

### Study Design and Participants

The Once-Weekly Semaglutide in People with HIV-Associated Lipohypertrophy study was a single-center, randomized, double-blind, placebo-controlled phase 2b trial (ClinicalTrials.gov NCT04019197) conducted at University Hospitals Cleveland Medical Center (Cleveland, OH). The primary trial objective was to evaluate semaglutide’s effects on body fat distribution in PLWH over 32 weeks. Key inclusion criteria were: age ≥18 years, documented HIV-1 infection on stable ART for ≥12 weeks, HIV-1 RNA <400 copies/mL for ≥6 months, body mass index (BMI) ≥25 kg/m², and clinical evidence of lipohypertrophy (central fat accumulation) with waist circumference >95 cm (men) or >94 cm (women) and waist-to-hip ratio >0.94 (men) or >0.88 (women). Participants with diabetes or active cardiovascular disease were excluded, as were those pregnant or with uncontrolled comorbidities. A total of 154 individuals were screened, 108 were randomized (54 to each group). Eight participants (15%) in each arm discontinued prematurely, leaving 92 who completed 32 weeks; of these, 84 had paired samples available for epigenetic analysis. Randomization was 1:1 to semaglutide or placebo, stratified by sex, using block sizes of six via an online randomization system. Both participants and investigators were blinded to treatment assignment. Semaglutide was administered by experienced clinic nurses subcutaneously once weekly, with dose escalation (0.25 mg for 4 weeks, to 0.5 mg for 4 weeks, then to 1.0 mg) then 1.0 mg weekly through week 32. The placebo group received volume-matched saline injections on the same schedule. The trial protocol was approved by the Institutional Review Board, and all participants provided written informed consent.

### Outcomes and Assessments

While the parent trial’s primary outcomes were changes in adipose tissue volume (measured by CT) and body composition (DXA scans) at 32 weeks, our current analysis focuses on epigenetic aging markers as secondary/exploratory outcomes. Peripheral blood mononuclear cells (PBMCs) were collected by phlebotomy at baseline and week 32 visits, isolated by Ficoll gradient, and cryopreserved at –80°C until analysis. Genomic DNA was extracted from thawed PBMC samples using Qiagen kits. Genome-wide DNA methylation profiling was performed using the Infinium MethylationEPICV2 BeadChip (Illumina), which covers >850,000 CpG sites, following manufacturer protocols. Briefly, 500 ng of genomic DNA per sample was bisulfite-converted (Zymo EZ DNA Methylation kit) and hybridized to the EPIC array, with arrays scanned on an iScan instrument to produce raw intensity data. To pre-process the methylation data, we used the *minfi* pipeline(Aryee et al. 2014), and low quality samples were identified using the *qcfilter()* function from the ENmix package(Xu et al. 2016), using default parameters. 100% of the original samples passed the QA/QC (p < 0.05) and were deemed to be high quality samples.

### Epigenetic Clock Calculations

We examined three generations of epigenetic clocks: first-generation clocks that estimate chronological age (Horvath1, Horvath2, and Hannum clocks); second-generation clocks that predict mortality/morbidity risk (PhenoAge and GrimAge, including a principal-component version of GrimAge denoted PCGrimAge); and a third-generation clock known as DunedinPACE, which measures the pace of aging (years aged per chronological year). To improve robustness for longitudinal analysis, we used principal component-based versions of the first- and second-generation clocks (denoted “PC clocks”) wherever applicable. These PC clocks leverage principal components of DNAm data associated with the original clock algorithms, enhancing technical reliability and reducing noise in repeated measures. Published epigenetic clocks were calculated according to published methods from processed DNA methylation data. To calculate the principal component-based epigenetic clock for the Horvath multi-tissue clock, Hannum clock, DNAmPhenoAge clock, GrimAge clock, and telomere length we used the custom R script available via GitHub (https://github.com/MorganLevineLab/PC-Clocks). Non-principal component-based (non-PC) Horvath, Hannum, and DNAmPhenoAge epigenetic metrics were calculated using the *methyAge* function in the ENMix R package. The pace of aging clock, DunedinPACE, was calculated using the *PACEProjector* function from the DunedinPACE package available via GitHub (https://github.com/danbelsky/DunedinPACE). We used a 12 cell immune deconvolution method to estimate cell type proportions(Zheng et al. 2018). For Biolearn, DNA methylation beta values (ssNoob-normalized) and matched sample metadata were imported into R (v4.3.2). Python integration was managed via the reticulate package, linking to a virtual environment with BioLearn installed. Missing CpGs were imputed using dataset-wide means via impute_from_average(), and the resulting matrix was combined with metadata into a GeoDataobject. The GrimAgeV1 and GrimAgeV2 models, obtained from the BioLearn ModelGallery, were applied using default parameters. Both models are based on Cox proportional hazards regression, trained to predict time-to-death from DNA methylation profiles. Internally, the models first extract a subset of CpGs relevant to DNAm surrogates for plasma proteins and smoking pack-years, followed by transformation through weighted linear combinations. These component predictors are then integrated into a multivariate Cox-PH model to estimate mortality risk, which is scaled to generate biological age equivalents.

### Statistical Analysis

We computed the annual rate of epigenetic age change for each participant on each clock as: (AgeWeek32−AgeBaseline)/(32 weeks/52 weeks, yielding a “years per year” change. For DunedinPACE, which is already a per-year rate, we computed the change between Week32 and baseline values (so a negative change indicates slowing of aging). Group comparisons of these rates were first assessed with Student’s t-tests (two-sided) for an initial view. The primary analysis used ANCOVA to estimate the effect of treatment (semaglutide vs. placebo) on epigenetic aging rate, adjusting for prespecified covariates. Our model for each aging measure included the baseline value of that measure (to adjust for regression to the mean), treatment group, age, sex, baseline BMI, baseline HsCRP, and baseline sCD163. We also ran alternative models replacing BMI with visceral fat mass, or HsCRP with IL-6 or sCD14, with consistent results (data not shown). Interaction terms between treatment and key baseline factors (e.g., baseline epigenetic age acceleration or baseline BMI) were explored to see if the treatment effect varied by these factors. Because the study is exploratory for epigenetic outcomes, we did not adjust p-values for multiple comparisons across the different clocks; instead, we focus on consistency of the pattern of results. All analyses were conducted in R (v4.2.2).

## Data Availability

The epigenetic data generated (DNAm beta values for EPIC arrays) and analyzed during this study will be made available in a public repository upon publication, with controlled access to protect participant privacy. Summary data for epigenetic clock measures and relevant clinical variables are provided in the Supplementary Information. The trial’s clinical data are available from the corresponding author on reasonable request, in accordance with institutional data sharing policies. Anonymized data will be shared by request from a qualified academic investigator for the sole purpose of replicating procedures and results presented in the article.

## Code Availability

Epigenetic clock algorithms were applied using publicly available code or R packages as referenced above.

## Author contributions

Conceptualization: MJC and GM

Methodology: MJC

Investigation: GAM, DL

Supervision: MJC and GAM

Writing—original draft: MJC

Writing—review & editing: GAM

